# METABOLIC PROFILE IN PLASMA AND CSF OF LEVODOPA-INDUCED DYSKINESIA OF PARKINSON’S DISEASE

**DOI:** 10.1101/2020.11.17.20233551

**Authors:** Bruno L. Santos-Lobato, Luiz Gustavo Gardinassi, Mariza Bortolanza, Ana Paula Ferranti Peti, Ângela V. Pimentel, Lúcia Helena Faccioli, Elaine A. Del-Bel, Vitor Tumas

## Abstract

**Background:** The existence of few biomarkers and the lack of a better understanding of the pathophysiology of levodopa-induced dyskinesia (LID) in Parkinson’s disease (PD) require new approaches, as the metabolomic analysis, for discoveries.

**Objectives:** We aimed to identify a metabolic profile associated with LID in patients with PD in an original cohort, and to confirm the results in an external cohort (BioFIND).

**Methods:** In the original cohort, plasma and CSF were collected from 20 healthy controls, 23 patients with PD without LID, and 24 patients with PD with LID. LC-MS/MS and metabolomics data analysis were used to perform untargeted metabolomics. Untargeted metabolomics data from the BioFIND cohort were analyzed.

**Results:** We identified a metabolic profile associated with LID in PD, composed of multiple metabolic pathways. In particular, the dysregulation of glycosphingolipids metabolic pathway was more related to LID and was strongly associated with the severity of dyskinetic movements. Further, bile acid biosynthesis and C21-steroid hormone biosynthesis metabolites simultaneously found in plasma and CSF have distinguished patients with LID from other participants. Levels of cortisol and cortisone were reduced in patients with PD and LID compared to patients with PD without LID. Data from the BioFIND cohort confirmed dysregulation in plasma metabolites from the bile acid biosynthesis and C21-steroid hormone biosynthesis pathways.

**Conclusion:** There is a distinct metabolic profile associated with LID in PD, both in plasma and CSF, which may be associated with the dysregulation of lipid metabolism and neuroinflammation.

## Introduction

Levodopa-induced dyskinesias (LID) are a major motor complication in Parkinson’s disease (PD) after chronic use of levodopa, occurring in 33 to 51.2% of patients with PD after 5 years of levodopa therapy [1] and causing increased health care costs and worsening of the quality of life [2–5]. Variables as age at PD onset and levodopa dose are important clinical risk factors associated with LID [6,7], as well as the association of some genetic polymorphisms [7]. Maladaptive plastic changes on the basal ganglia neurotransmission [8] and neuroinflammation [9–14] are the main processes associated with LID pathophysiology.

The development of biomarkers in neurological diseases is essential for a better understanding of its pathophysiology, but there are few biomarkers associated with LID in PD. For the discovery of a possible signature and molecular pathogenesis of LID in PD, untargeted metabolomics may be a strategic initial approach concerned with the identification and quantification of metabolites and metabolic pathways linked to a specific condition [15].

Previous studies have mostly explored metabolomics in blood and CSF of patients with PD, and a broad number of metabolic pathway dysfunctions have been associated with the disease [15–18]. Also, PD progression could be estimated based on a panel of metabolites discovered by metabolomic analysis [19], and no difference was seen in the metabolic profile of genetic forms of PD [20]. About metabolomic studies in LID, a previous study showed no association between dyskinesias and the kynurenine pathway in plasma [21], even though another study reported a specific metabolite signature based on kynurenine products in plasma and CSF on LID [22].

To explore metabolites as possible biomarkers of LID, we conducted a study to identify a metabolite profile in plasma and CSF associated with LID in patients with PD. After, we explored the metabolite profile in plasma and CSF associated with LID in another cohort (BioFIND cohort).

## Methods

### Study design and participants

We conducted an observational cross-sectional study to analyze the metabolic profile in plasma and CSF of patients with PD with and without LID and in healthy controls. For more details about the enrollment of participants, inclusion and exclusion criteria, and clinical evaluations, see Marchioni et al. [23]. For evaluation, we collected clinical and epidemiological data, Hoehn and Yahr stage, the MDS-UPDRS, and the Unified Dyskinesia Rating Scale (UDysRS). For analysis, participants were divided into three groups: healthy controls (HC), patients with PD without LID (PD-ND), and patients with PD with LID (PD-D). The study was approved by the Ribeirão Preto Medical School Ethics Committee (Number 3.036.243), and all participants provided written informed consent.

Additionally, we used the database from the Fox Investigation for New Discovery of Biomarkers in Parkinson’s Disease (“BioFIND”) study cohort (http://biofind.loni.usc.edu/), a cross-sectional and observational study of moderate to advanced patients with PD and healthy controls, conceived for biomarker discovery in PD [24]. For up-to-date information on the study, visit www.michaeljfox.org/biospecimens.

### Collection, processing, and storage of biologic samples

As previously described [23], all patients were instructed to take their regular morning dose of dopaminergic drugs, and we collected peripheral blood by venipuncture and CSF through lumbar puncture on the same day. After, whole blood was centrifuged at 4°C and 1600×g for 15 minutes; CSF was gently mixed to avoid gradient effects and centrifuged at 4°C at 4000×g for 10 minutes. Supernatant plasma from whole blood and centrifuged CSF samples were aliquoted into 1-mL cryotubes and stored at -80°C until use. In the BioFIND cohort, blood plasma samples and CSF samples were also collected and immediately frozen in a –80°C freezer [24].

### Sample preparation for untargeted metabolomic analysis and metabolite quantification by LC-MS/MS

Samples were extracted using solid-phase extraction (SPE) as described previously [25] for the extraction of eicosanoids in human plasma. The samples were spiked with 10 μL internal standards (IS) (corticosterone-d_4_ at 50 ng·mL^−1^ in methanol). Following the extraction, the samples were dried, suspended in 50 μL of methanol, and injected into the LC-MS/MS system. Before the extraction process, 300 μL of plasma samples with IS were denatured with 1.5 mL of methanol/acetonitrile (1:1, v/v) at 4°C. The denatured proteins were removed by centrifugation (400×g, 20 min), and the supernatants were diluted in water to decrease the methanol/acetonitrile concentration to 10%. SPE was using C18 SPE cartridges, and the first step consisted of conditioning the cartridge with 2 mL of methanol and 2 mL of an aqueous solution of acetic acid 0.1%. After, the samples were loaded, and 2 mL of an aqueous solution of acetic acid 0.1% was used for the removal of impurities. For sample elution were used 2 mL of methanolic solution of acetic acid 0.1%

### Metabolomics data analysis

Untargeted metabolomics data was performed with full scan mass spectral data acquired in positive mode, with mass-to-charge ratio (*m/z*) ranging from 100 - 1500, as previously described [26]. The package *apLCMS* [27] was used for peak peaking, noise filtering, retention time and m/z alignment, and feature quantification. A metabolite feature is characterized by three parameters: *m/z*, retention time, and intensity. Data were log2 transformed and normalized by the mean. Only features detected in 90% of plasma (4423 metabolite features) and CSF (4259 metabolite features) were retained for statistical analyses. Missing values were imputed by half mean of a particular feature across all samples. The *mummichog* software v1.0.10 was used for metabolic pathway enrichment analysis (mass accuracy under 10 ppm) [28].

Untargeted metabolomics data from the BioFIND study cohort was acquired and processed by the company Metabolon Inc. Scaled and median normalized data were log2 transformed and used for statistical analysis.

### LC-MS/MS and MRM^HR^ analysis for quantification of plasma and CSF cortisol and cortisone levels

To confirm dysregulation of the C21-steroid hormone biosynthesis pathway through two metabolites (cortisol and cortisone), LC-MS/MS analysis was performed according to the methodology described by Peti et al. [29]. In summary, the LC-MS/MS was carried out on a Nexera Ultra-High Performance Liquid Chromatographic (UHPLC) system by Shimadzu (Kyoto, HO, JP) and a TripleTOF® 5600+ mass spectrometry from AB Sciex (Foster, CA, USA) equipped with a Turbo-V IonSpray and Calibrant Delivery System (CDS), operated in positive mode. Mobile phases used for chromatographic separation were formic acid 0.1% (A) and acetonitrile, containing 0.1% formic acid in both. The column was an Ascentis Express C18 (100 x 2.1 mm; 2.7 µm) from Supelco (St. Louis, MO, USA). A flow rate of 0.5 mL·min^-1^ and an injection volume of 10 μL were used. The gradient condition was described in Appendix S1 (Table S1). The optimized MS parameters were as follows: gas1: 60 psi; gas2: 40 psi; curtain gas: 25 psi; ion spray voltage: 5 kV; turbo temperature: 550 °C; *m/z* range: 50 a 400; and dwell time: 100 ms.

MRM^HR^ transition used for cortisol and cortisone detection were 363.2166> 267.1740 and 361.2010> 163.1115, respectively. MultiQuant™ 3.0.2 software (Foster, CA, USA) was used for data analysis.

### Statistical analysis

To compare two independent groups, we used the Mann-Whitney test; to compare three or more groups, we performed the Kruskal-Wallis test for continuous variables (followed by Dunn’s test for multiple comparisons), and the chi-square test for categorical variables. To compare two continuous variables, we performed Spearman’s Rho correlation test. Analyses were performed using SPSS for Windows version 23.0 (SPSS Inc., Chicago, USA), and figures were made using GraphPrism for Windows version 5.0 (GraphPad Software Inc., La Jolla, USA). Also, part of these statistical analyses and visualization of untargeted metabolomics data were performed using the software environment for statistical computing and graphics R v3.4.0 and associated packages such as *limma* (to calculate differential abundance), *gplots* (to generate heat maps), *amap* (to perform hierarchical clustering), *ggplot2* (to generate Manhattan and Bubble plots).

## Results

### Clinical and epidemiological data

In the original cohort, we recruited a total of 67 participants who fulfilled the inclusion and exclusion criteria. We did not perform CSF analysis in 4 patients with PD (with LID – 1 patient, without LID – 3 patients) due to technical problems with lumbar puncture. Data are summarized in Appendix S1 (Table S2).

### Plasma and CSF metabolite profile associated with PD and LID

Our untargeted metabolomics analysis detected over 7000 metabolite features, of which 4423 detected in the plasma and 4259 detected in the CSF were used for statistical analysis. In plasma, 249 metabolites (subset 1) distinguished between PD-D and other groups (PD-ND and HC) (Figure 1A), and in CSF, 382 metabolites (subset 2) distinguished between PD-D and other groups (Figure 1B). These subsets of metabolites provided good differentiation between patients with PD and LID and other groups, both in plasma and in CSF (Figure 1C and 1D). Furthermore, 104 metabolites (subset 3) were detected both in plasma and CSF distinguished between PD-D and other groups.

**Figure 1.**
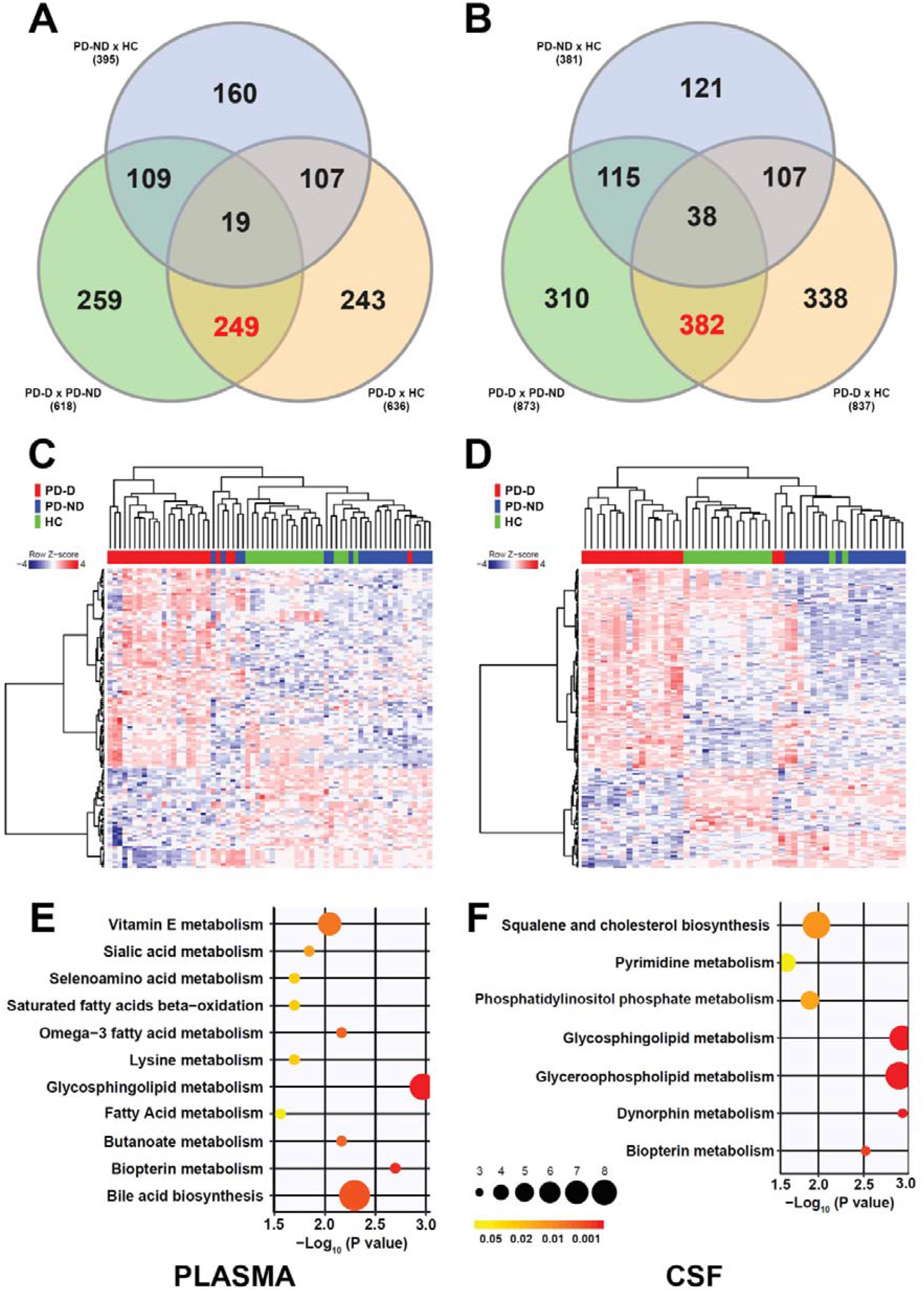
Metabolomic analysis of patients with PD with LID. A and B: Venn diagrams of differentially abundant metabolite features between patients with PD and LID (PD-D), patients with PD without LID (PD-ND) and healthy controls (HC), detected in plasma (A) and CSF (B) datasets. Significant features were selected by the moderated t-test with *limma* package for R. Red values indicate the number of significant metabolites overlapping in comparisons between PD-D vs PD-ND and PD-D vs HC, but not between PD-D vs HC. C and D: Heat maps showing two-way hierarchical clustering based on the intensity of highly significant metabolite features differing between groups in (C) plasma (135 *m/z* features, p < 0.001, FDR < 0.035) and (D) CSF (186 *m/z* features, p < 0.001, FDR < 0.025). Significant features were selected by ANOVA with *limma* package for R. The blue-to-red scale indicates lower to higher intensity levels based on a Z-score of normalized intensities for each metabolite feature. E and F: Metabolic pathway analysis showing enriched metabolic pathways in plasma (E) and CSF (F). The color of the circles indicates the significance of the pathway, and the size of the circles indicate the number of metabolites involved in the pathway.

### Metabolic pathways associated with PD and LID

Using the subsets 1 (plasma) and 2 (CSF) of metabolites associated with LID in PD, we detected dysfunctions in multiple metabolic pathways. In plasma, there were dysfunctions in 11 metabolic pathways: glycosphingolipids metabolism, biopterin metabolism, bile acid biosynthesis, omega-3 fatty acid metabolism, butanoate metabolism, vitamin E metabolism, sialic acid metabolism, selenoamino acid metabolism, saturated fatty acid beta-oxidation, lysine metabolism, and fatty acid metabolism, in decrescent order of significance (Figure 1E). In CSF, there were dysfunctions in 7 metabolic pathways: glycosphingolipids metabolism, glycerophospholipids metabolism, dynorphin metabolism, biopterin metabolism, squalene and cholesterol biosynthesis, phosphatidylinositol phosphate metabolism, and pyrimidine metabolism, in decrescent order of significance (Figure 1F). Considering the metabolites associated with LID in PD in both plasma and CSF (subset 3), we detected dysfunctions in 2 metabolic pathways: bile acid biosynthesis and C21-steroid hormone biosynthesis and metabolism (Figure 2). Metabolic pathways and metabolites associated with LID in PD are described in Appendix S1 (Table S3).

**Figure 2.**
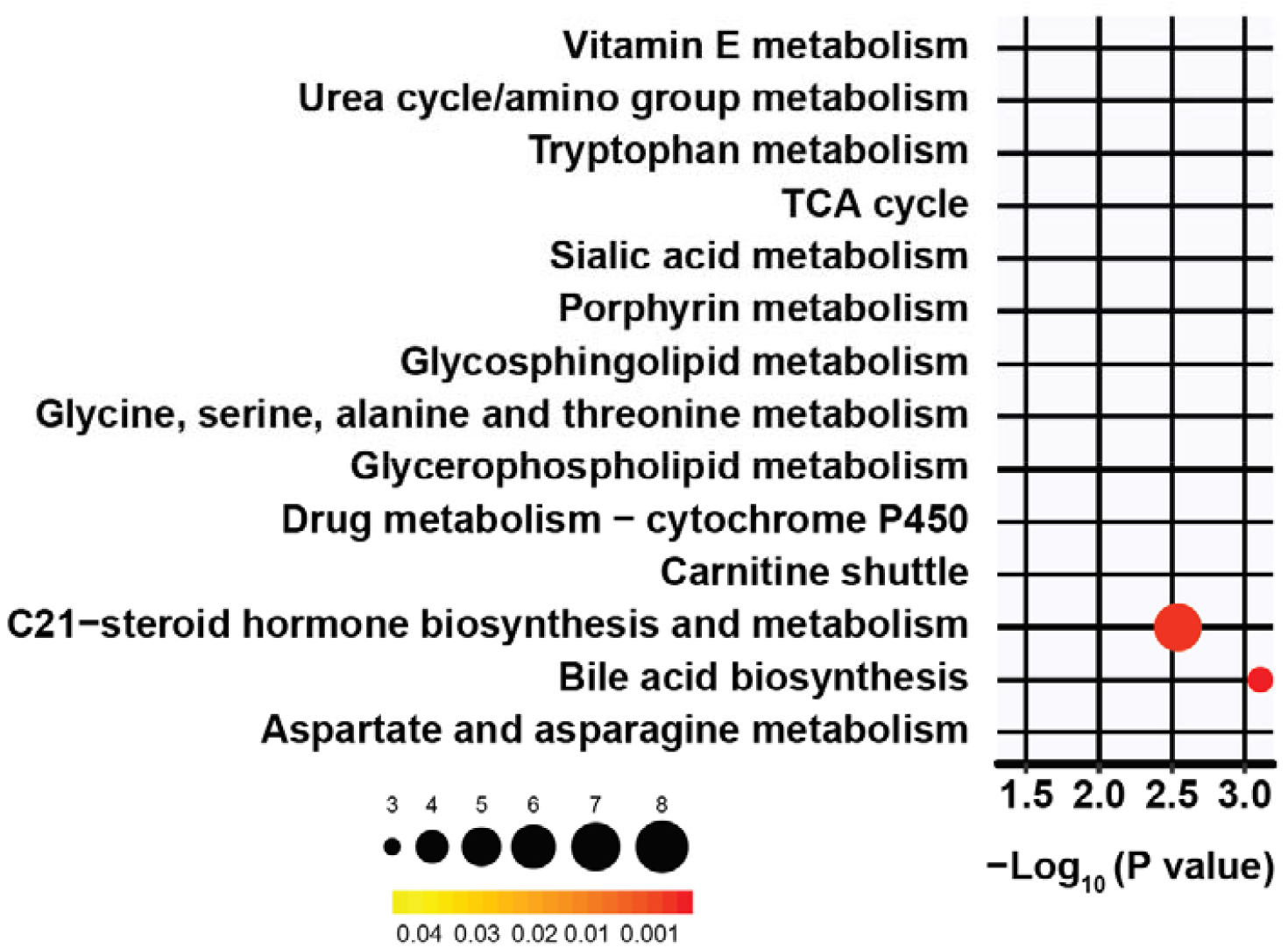
Metabolic pathway analysis showing enriched pathways in patients with PD and LID in metabolites found both in plasma and CSF. The color of the circles indicates significance of the pathway, and the size of the circles indicate the number of metabolites involved in the pathway.

### LID severity is associated with specific metabolic pathways

To evaluate the association between plasma and CSF metabolites with LID severity, we performed linear regression models adjusted by age, gender, disease duration, and LEDD using UDysRS as a continuous variable and marker of LID severity. After, we used the p-values extracted from linear regression models to identify the main biochemical pathways associated with LID severity using the *mummichog* software [28]. There was a match in some metabolic pathways associated with LID by hierarchical clustering analysis and linear regression model in plasma (vitamin E metabolism) and CSF (pyrimidine metabolism, glycosphingolipids metabolism, glycerophospholipids metabolism) (Table 1 and Figure S1 in Appendix S1).

**Table 1.**
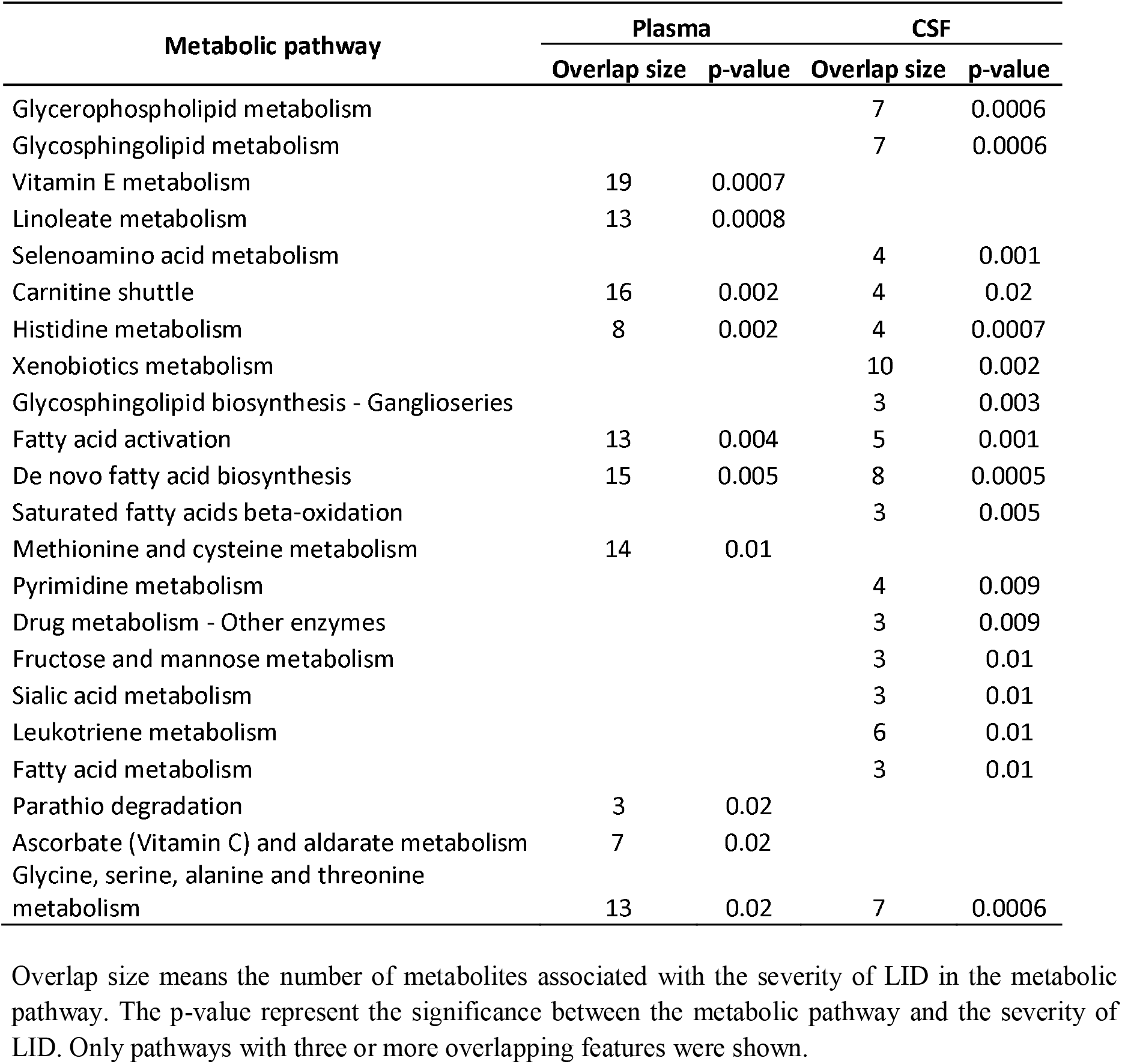
Association between metabolic pathways and severity of LID in plasma and CSF.

### Metabolites from bile acid biosynthesis and C21-steroid hormone biosynthesis pathways are associated with LID in the BioFIND cohort

We included BioFIND cohort data from 41 patients with PD (with LID – n = 21; without LID – n = 20) for analysis of plasma metabolites, and 44 patients with PD (with LID – n = 22; without LID – n = 22) for analysis of CSF metabolites. For both plasma and CSF samples, 772 metabolites were quantified. Relative metabolite abundances (after normalization) were compared between plasma and CSF samples from patients with and without PD, and only 4 metabolites in plasma showed differences between groups: deoxycholate, taurodeoxycholate, 3β-7α-dihydroxy-cholestenoate (bile acid biosynthesis pathway), and corticosterone (C21-steroid hormone biosynthesis pathway) (Figure 3).

**Figure 3.**
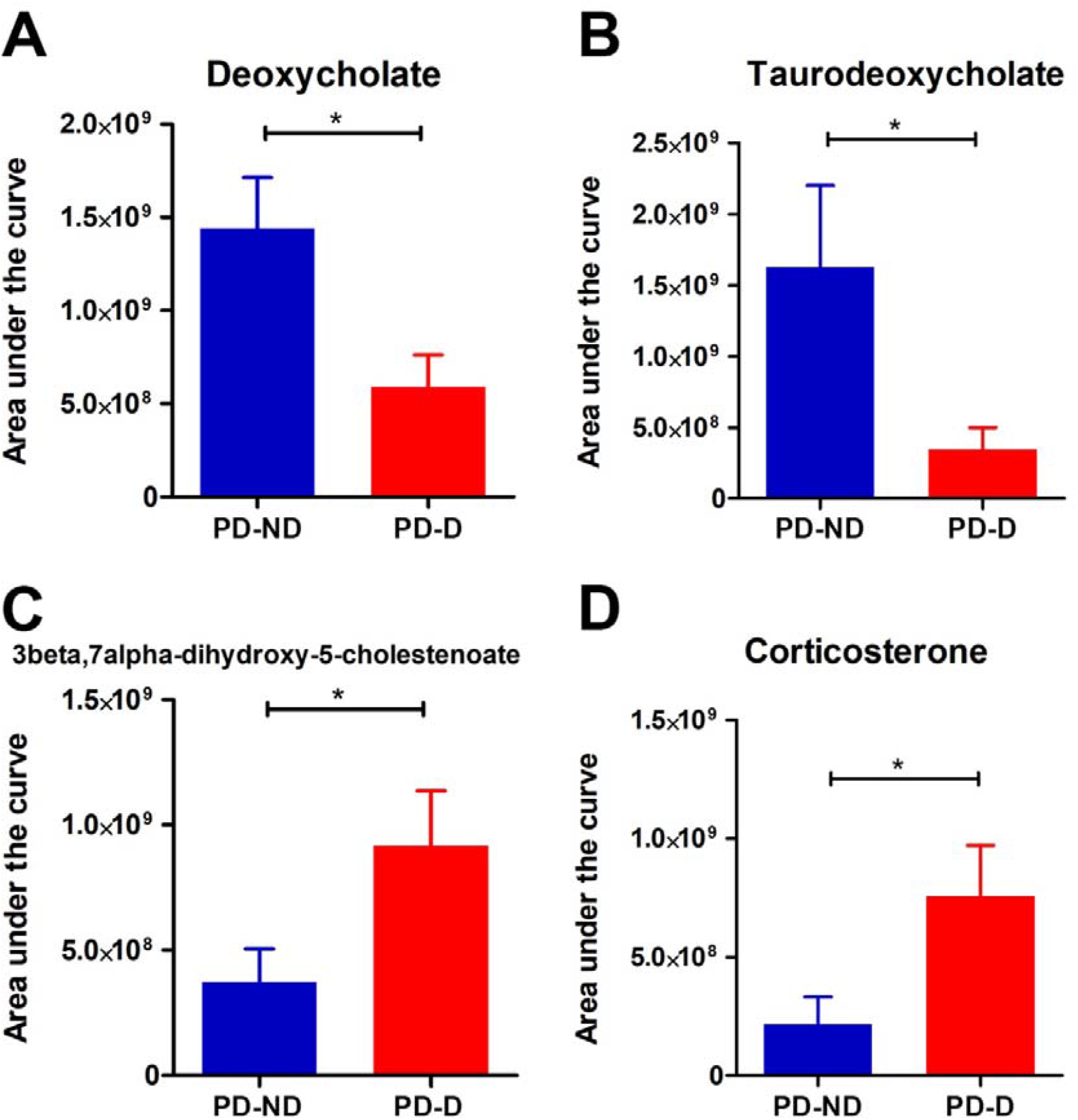
Relative abundance of top four metabolites in plasma from participants of BioFIND cohort, according to patients with PD with and without LID. A: Deoxycholate. B: Taurodeoxycholate. C: 3β,7α-dihydroxy-5-cholestenoate. D: Corticosterone.

### Plasma and CSF cortisol and cortisone levels in patients with PD and LID

Aiming to support the association of C21-steroid hormone biosynthesis pathway with LID, we compared plasma and CSF levels of cortisol and cortisone between PD-D, PD-ND and HC groups. Patients with PD and LID had lower plasma and CSF cortisol levels than patients with PD without LID (plasma: ANOVA F_2,63_ = 3.41, p = 0.03; PD-D versus PD-ND, Bonferroni mean difference = 565.4, p < 0.05; CSF: F_2,63_ = 4.96, p = 0.01; PD-D versus PD-ND, Bonferroni mean difference = 15.9, p < 0.05) (Figure 4). Also, patients with PD and LID had lower CSF cortisone levels than patients with PD without LID (CSF: F_2,63_ = 9.61, p = 0.0002; PD-D versus PD-ND, Bonferroni mean difference = 0.35, p < 0.05) (Figure 4).

**Figure 4.**
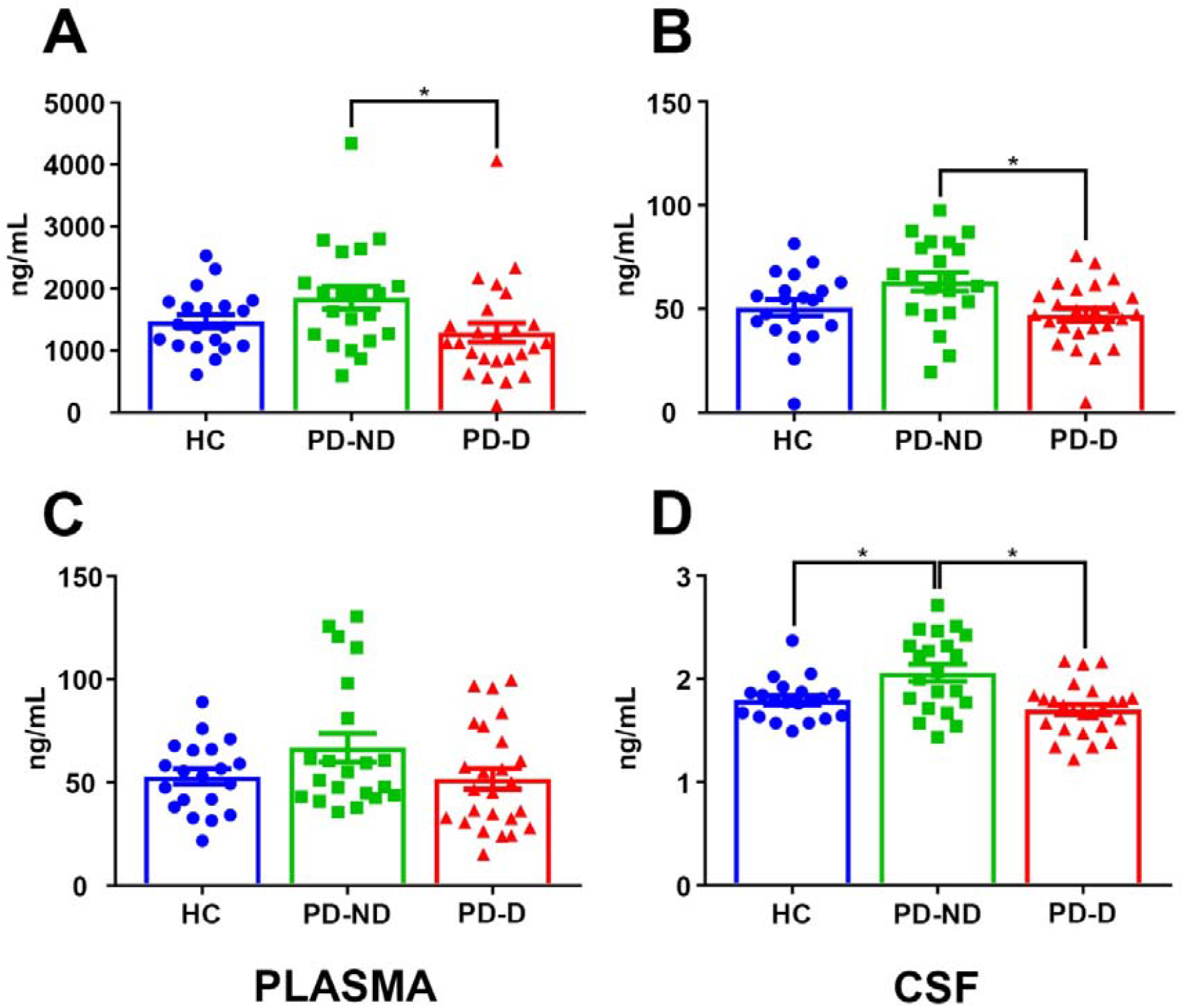
Plasma and CSF cortisol and cortisone levels in patients with PD and healthy controls. A and B: Cortisol. C and D: Cortisone.

Overall, we found a weak positive correlation between plasma and CSF cortisol levels (r = 0.4, p = 0.001, n = 63) and a moderate positive correlation between plasma and CSF cortisone levels (r = 0.53, p < 0.001, n = 63). There were no correlations between clinical variables (age at PD onset, weight, disease duration, levodopa therapy duration, LEDD, MDS-UPDRS, Hoehn and Yahr stage, and UDysRS) and plasma and CSF cortisol and cortisone levels.

## Discussion

Our results showed that there are metabolite profiles associated with LID in PD in segregated subsets of metabolites in plasma and CSF. These metabolite profiles comprise multiple metabolic pathways (in plasma: glycosphingolipid metabolism, biopterin metabolism, bile acid biosynthesis, omega-3 fatty acid metabolism, butanoate metabolism, vitamin E metabolism, sialic acid metabolism, selenoamino acid metabolism, saturated fatty acid beta-oxidation, lysine metabolism, and fatty acid metabolism; in CSF: glycosphingolipid metabolism, glycerophospholipid metabolism, dynorphin metabolism, biopterin metabolism, squalene and cholesterol biosynthesis, phosphatidylinositol phosphate metabolism, and pyrimidine metabolism). Glycosphingolipid metabolism was the metabolic pathway more dysregulated in patients with PD with LID. And between these metabolic profiles associated with LID, glycosphingolipid metabolism and glycerophospholipid metabolism pathways in CSF were strongly associated with the severity of LID symptoms in patients with PD, together with the vitamin E metabolism in plasma.

In the subset of metabolites found simultaneously in plasma and CSF, there was also a metabolic profile associated with LID: dysregulation in bile acid biosynthesis and C21-steroid hormone biosynthesis and metabolism pathways were able to discriminate patients with PD with LID from other groups. Also, plasma metabolites belonging to bile acid biosynthesis and C21-steroid hormone biosynthesis pathways were associated with LID in an independent cohort (BioFIND). We measured the levels of two metabolites of the C21-steroid hormone biosynthesis pathway (cortisol and cortisone): CSF cortisol and cortisone levels were lower in patients with PD and LID than in patients with PD without LID.

The association of glycosphingolipid and LID in PD has not been widely explored. In a rat model of LID, sphingolipid metabolism and signaling pathways were dysregulated in the striatum [30]. Also, growing evidence shows the role of sphingolipids/glycosphingolipids in neuroinflammation through its interaction with microglia, chemical mediators of inflammation (cytokines, chemokines), and inflammation-associated signaling pathways [31,32].

In humans, there is no clear association between LID and sphingolipids/glycosphingolipids metabolism, despite some evidence showing the sphingolipid metabolic pathway is enriched in the plasma of patients with PD [17]. The activity of important enzymes associated with the metabolism of glycosphingolipids (glucocerebrosidase, alpha-galactosidase A, beta-galactosidase, beta-hexosaminidase) is dysregulated in patients with PD [32]. Furthermore, there are many disorders of sphingolipids/glycosphingolipids metabolism identified as lysosomal storage diseases, as Niemann-Pick disease, Gaucher’s disease, Farber’s disease, and Krabbe’s disease [33]. Gaucher’s disease, characterized by the absence of beta-glucocerebrosidase (GBA) enzyme and accumulation of glucosylceramide and glucosylsphingosine, has been linked with PD in the last decades due to the discovery of mutations in *GBA1* as the main risk factor for PD [34], and patients with PD and heterozygous *GBA1* mutations present earlier disease onset, increased cognitive impairment and more frequent pain [34]. Patients with PD and *GBA1* mutations may have a higher risk of developing LID [35,36], but there are also inconclusive results [34]. For these patients, ambroxol therapy may increase GBA activity, and a recent clinical trial reported a slight reduction in motor symptoms after six months of ambroxol in patients with PD and *GBA1* mutations, but unfortunately motor complications, as LID, were not evaluated [37].

As an antioxidant, vitamin E was explored in PD clinical trials as a possible neuroprotective therapy against the oxidative stress on its pathophysiology, including the well-known DATATOP (Deprenyl and Tocopherol Antioxidative Therapy of Parkinsonism) study; no study showed an effect on preventing or delaying the progression of the disease [38].

Notwithstanding the absence of association between vitamin E metabolism pathway and LID, studies on tardive dyskinesia, a complication of chronic neuroleptic therapy which causes abnormal involuntary movements similar to LID, reported reduction of involuntary movements with the supplementation of vitamin E [38].

There is no direct evidence associating a possible role of bile acids on LID in PD. Previous studies showed bile acids (tauroursodeoxycholic acid, ursodeoxycholic acid) may increase the survival of dopaminergic nigral cells after transplantation through anti-apoptotic mechanisms [39] and reverse mitochondrial dysfunction in *LRRK2* mutation carriers [40]. As sphingolipids/glycosphingolipids, bile acids are also implicated in neuroinflammation as antiinflammatory agents, reducing astrocyte activation and expression of interferon-1-beta in a mouse model of PD [41], which may have implications for the pathophysiology of LID in PD.

In patients with PD, a metabolomic study in plasma showed the bile acid biosynthesis pathway was dysregulated in patients with PD, together with other lipid metabolic pathways (glycerolphospholipids, fatty acids), and considering the essential role of bile acids on lipid homeostasis [42], the authors suggested that abnormalities in bile acid may cause disturbances in all lipid metabolism [43]. These findings were supported by another recent study, which reported a significant metabolic change in the bile acids pathway, both in a murine model of PD and in patients with PD [20]. Also, another study proposed that abnormalities in bile acids biosynthesis in patients with PD caused by intestinal dysbiosis, which is common in PD, may dysregulate lipid metabolism [44]. Cerebrotendinous xanthomatosis, a rare inborn error of cholesterol and bile acid metabolism associated with parkinsonism, is caused by the deficiency of sterol 27-hydroxylase (CYP27), leading to the absence of chenodeoxycholate, used in secondary bile acid synthesis, and resulting in deposits of cholesterol and cholestanol in tissues, including the brain [45].

The interaction of steroid hormones with dopaminergic neurotransmission stimulated many animal studies using LID models. Estradiol, corticosterone, and dehydroepiandrosterone showed a consistent antidyskinetic effect on parkinsonian animals with chronic levodopa therapy [46–48]. The administration of finasteride, a 5-alpha reductase inhibitor which blocks the conversion of testosterone into dihydrotestosterone, also reduced dyskinetic movement in a LID animal model [49]. Neuroinflammation has been seen as a main mechanism underlying the steroid-mediated reduction of LID [48]. In patients with PD, a double-blind randomized clinical trial of low-dose estrogen replacement therapy showed the same frequency of LID in post-menopausal women with PD using estrogen or not using [50]. Regarding the association between the C21-steroid hormone biosynthesis pathway and PD, a metabolomic study showed a decrease in testosterone levels in the serum of patients with PD [18].

Together, most of these metabolic pathways dysregulated in dyskinetic patients with PD in this study are part of the metabolism of lipids (glycosphingolipids, bile acid biosynthesis, C21-steroid hormone biosynthesis). Considering the role of lipids in neuroinflammation [51], the dysregulation of the lipid metabolism in the plasma and CSF of patients with PD and LID support the hypothesis that a chronic pro-inflammatory state in the brain leads to LID.

As a limitation, we did not include a drug-näive group of patients with PD, which would allow us to investigate variations in the metabolic profile of untreated patients.

In conclusion, these findings suggest that there is a distinct metabolic profile associated with LID in PD, both in plasma and CSF, which may be associated with the dysregulation of lipid metabolism and neuroinflammation.

## Data Availability

The datasets generated during and/or analyzed during the current study are available from the
corresponding author on reasonable request.

## Acknowledgments

We would like to thank Manuelina Macruz Capelari, Nathália Novaretti, and Larissa Serveli for technical support.

Parts of the data used in the preparation of this article were obtained from the Fox Investigation for New Discovery of Biomarkers (“BioFIND”) database (http://biofind.loni.usc.edu/). For up-to-date information on the study, visit www.michaelifox.org/biofind. BioFIND is sponsored by The Michael J. Fox Foundation for Parkinson’s Research (MJFF) with support from the National Institute for Neurological Disorders and Stroke (NINDS).

## Author’s Role

BLSL: 1A, 1B, 1C, 2A, 2B, 2C, 3A, 3B

LGG: 1C, 2A, 2B, 2C, 3B

MB: 1C, 2B, 2C, 3B

APFP: 1C, 3B

AVP: 1C, 3B

LHF: 1C, 2C, 3B

EADB: 1A, 1B, 2C, 3B

VT: 1A, 1B, 2C, 3B

---

**Supplement Table 1.** Chromatographic gradient used for LC-MS/MS analysis.

| Time (min) | Mobile Phase A (%) | Mobile Phase B (%) |
| --- | --- | --- |
| 0 | 70 | 30 |
| 1 | 70 | 30 |
| 2 | 50 | 50 |
| 5 | 40 | 60 |
| 8 | 40 | 60 |
| 9 | 02 | 98 |
| 11 | 02 | 98 |

**Supplement Table 2.** Clinical and epidemiological data of patients with Parkinson’s disease and healthy controls.

| General characteristics | HC (n=20) | PD-ND (n=20) | PD-D (n=23) | p-value |
| --- | --- | --- | --- | --- |
| Male sex, % (n) | 30 (6) | 75 (15) | 65.2 (15) | 0.002<sup>a</sup> |
| Age at the time of evaluation<sup>b</sup> | 63 (61-67) | 62 (53-66) | 59 (53-64) | 0.06<sup>c</sup> |
| Weight (kg) | NA | 72.5 (66-79) | 73 (60-83) | 0.63<sup>d</sup> |
| Age at onset of PD<sup>b</sup> | NA | 54.5 (48-61) | 47 (42-51) | 0.03<sup>d</sup> |
| Disease duration (years)<sup>b</sup> | NA | 8 (5-12) | 11 (8-16) | 0.01<sup>d</sup> |
| Levodopa therapy duration (years)<sup>b</sup> | NA | 5 (3-8) | 8 (6-12) | 0.05<sup>d</sup> |
| Levodopa dose at evaluation (mg/day)<sup>b</sup> | NA | 500 (400-850) | 850 (750-1200) | 0.003<sup>d</sup> |
| Levodopa equivalent daily dose (mg/day)<sup>b</sup> | NA | 950 (500-1275) | 1400 (1090-2050) | 0.002<sup>d</sup> |
| Tremor as initial motor symptom, % (n) | NA | 70 (14) | 43.5 (10) | 0.12<sup>a</sup> |
| Tremor dominant phenotype at evaluation, % (n) | NA | 80 (16) | 26.1 (6) | 0.002<sup>a</sup> |
| MDS-UPDRS Part I<sup>b</sup> | NA | 10.5 (3-17) | 10 (8-17) | 0.28<sup>d</sup> |
| MDS-UPDRS Part II<sup>b</sup> | NA | 15 (5-18) | 19 (12-25) | 0.02<sup>d</sup> |
| MDS-UPDRS Part III<sup>b</sup> | NA | 30.5 (25-40) | 25 (19-34) | 0.3<sup>d</sup> |
| MDS-UPDRS Part IV<sup>b</sup> | NA | 0 (0-5) | 11 (8-15) | < 0.001<sup>d</sup> |
| MDS-UPDRS Total score<sup>b</sup> | NA | 57.5 (45-73) | 64 (57-88) | 0.04<sup>d</sup> |
| Hoehn & Yahr stage<sup>b</sup> | NA | 2 (2-2) | 2 (2-3) | 0.37<sup>d</sup> |
| UDysRS Part 1B<sup>b</sup> | NA | NA | 21 (16-25) | - |
| UDysRS Part 3<sup>b</sup> | NA | NA | 14 (11-19) | - |
| UDysRS Part 4<sup>b</sup> | NA | NA | 7 (4-9) | - |
| UDysRS Historical subscore<sup>b</sup> | NA | NA | 28 (17-39) | - |
| UDysRS Objective subscore<sup>b</sup> | NA | NA | 20 (15-27) | - |
| UDysRS Total score<sup>b</sup> | NA | NA | 53 (36-62) | - |

<sup>a</sup>Chi-square test comparing frequencies between healthy controls, patients with PD with and without levodopa-induced dyskinesias
<sup>b</sup>Values in median (interquartile range)
<sup>c</sup>Kruskal-Wallis test comparing medians between healthy controls, patients with PD with and without levodopa-induced dyskinesias
<sup>d</sup>Mann-Whitney test comparing medians between patients with and without levodopa-induced dyskinesias
Abbreviations: HC, healthy controls; NA, not applied; PD-D, Patients with PD with levodopa-induced dyskinesias; PD-ND, Patients with PD without levodopa-induced dyskinesias.

**Supplement Table 3.**
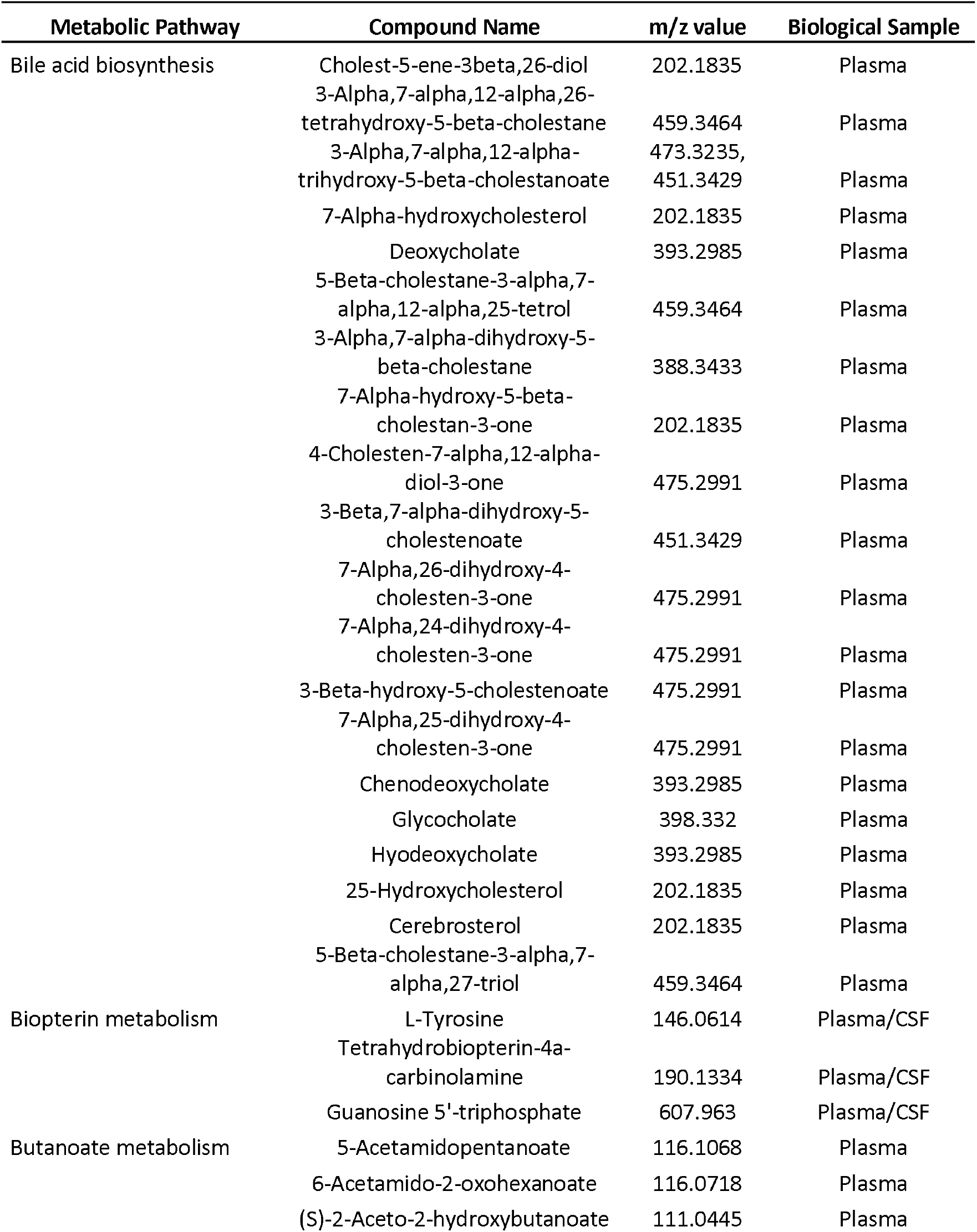
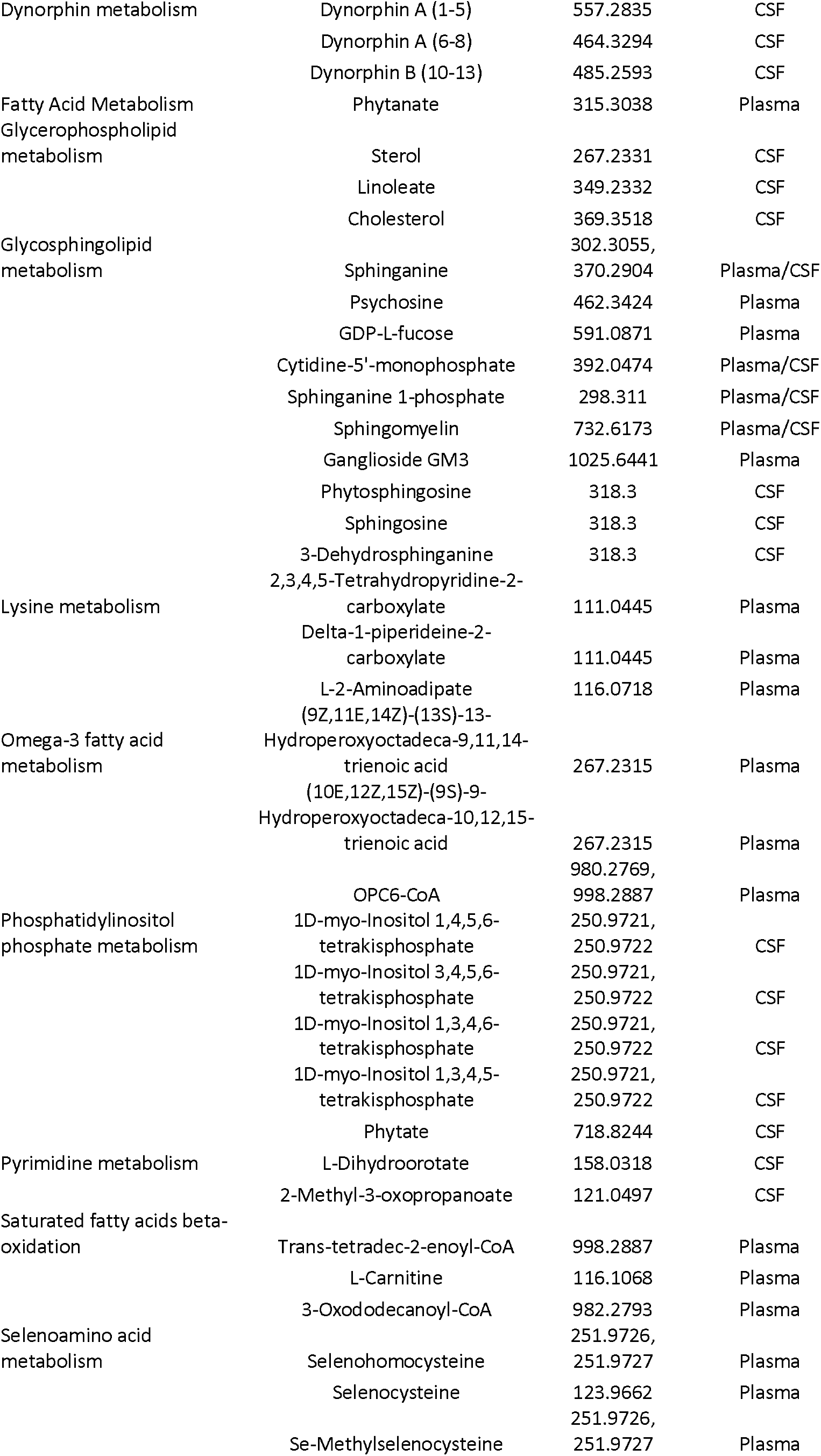
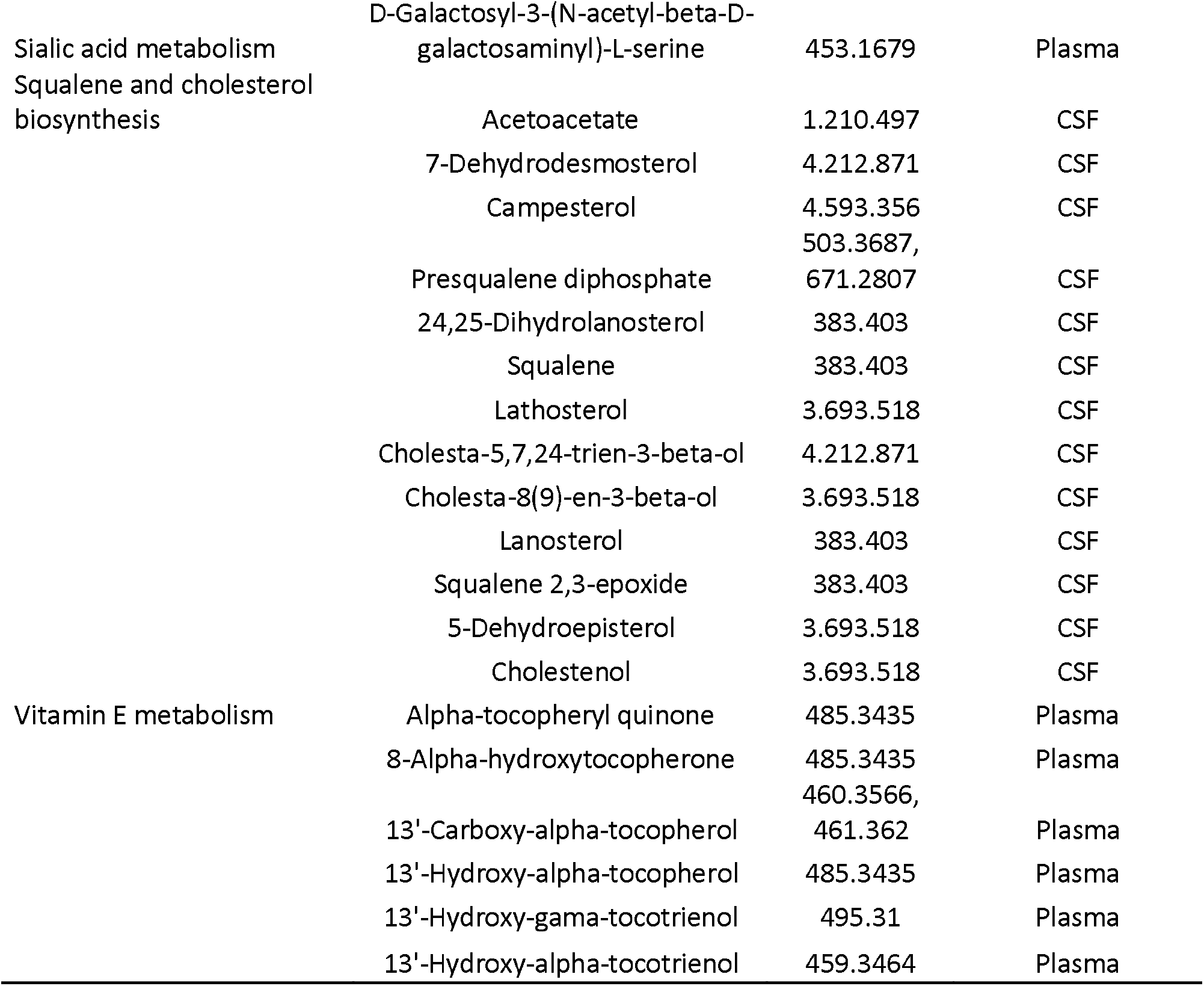
Metabolic pathways and metabolites associated with LID in PD, in plasma and CSF.

**Supplementary Figure 1.**
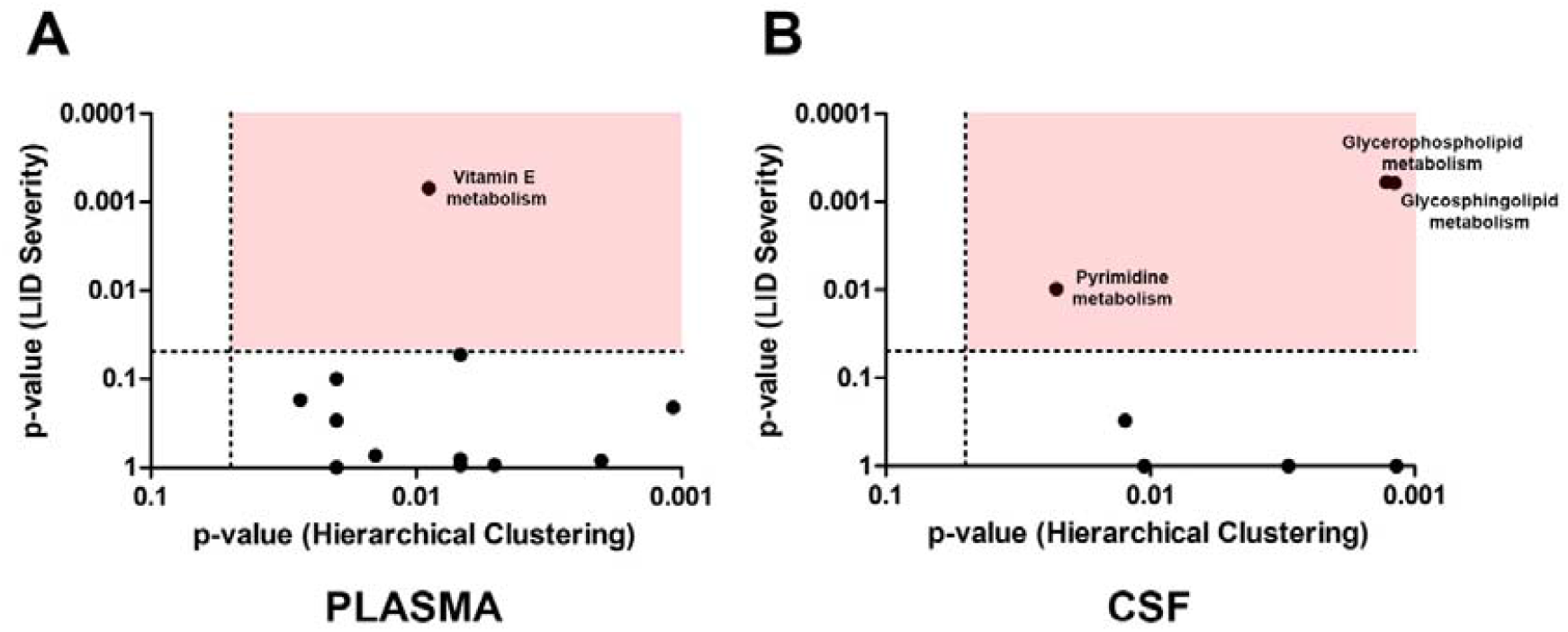
Correlations between p-values of the association of metabolic pathways with LID presence and the association of metabolic pathways with LID severity. A: Plasma. B: CSF. The association of metabolic pathways with LID presence was analyzed by hierarchical clustering, and the association of metabolic pathways with LID severity was analyzed by linear regression. Each black dot represents a metabolic pathway. Metabolic pathways more associated with both LID presence and dyskinesia severity are shown in the pink quadrant. Dotted lines in both axes indicate p-value = 0.05.

